# CARDIAC DYSFUNCTION DURING ADVERSE MATERNAL OUTCOMES IN HYPERTENSIVE DISORDERS OF PREGNANCY

**DOI:** 10.1101/2025.01.02.25319925

**Authors:** Veronica Giorgione, Jamie Kitt, Paul Leeson, Asma Khalil, Jamie O’Driscoll, Basky Thilaganathan

## Abstract

**Background:** Hypertensive disorders of pregnancy (HDP) are associated with significant cardiac remodeling during pregnancy and are important contributors to maternal morbidity and mortality. Whether acute adverse outcomes during HDP are associated with additional clinically relevant cardiac impairment has not been widely studied.

**Methods:** A prospective observational study was conducted on 255 women with HDP who underwent transthoracic echocardiography during the peripartum period. Maternal echocardiographic parameters, including left ventricular morphology and function, were analyzed to determine their association with adverse maternal outcomes by univariate and multivariate analyses. The composite adverse maternal outcome was defined as at least one of the following: admission to a high dependence unit, acute renal injury, adverse cardio-pulmonary events, stroke and disseminated intravascular coagulation.

**Results:** Adverse maternal outcomes occurred in 68 (26.7%) participants. Women with adverse outcomes had significantly higher left atrial volume index (LAVI) (28.8 [23.4-32.3] ml/m^2^ vs 26.6 [22.2-30.9] ml/m^2^, p=0.045) and E/e′ ratio (7.8 [6.6-9.2] vs 7.0 [5.9-8.1], p=0.002) compared to those without complications. In multivariable analysis, both LAVI (adjusted OR 1.09 [1.02-1.16], p=0.009) and E/e′ ratio (adjusted OR 1.25 [1.04-1.49], p=0.018) remained independently associated with adverse maternal outcomes after adjusting for maternal factors and clinical variables.

**Conclusions:** Cardiac abnormalities, particularly in diastolic function, are more common in women with adverse maternal outcomes in HDP. Whether enhanced cardiovascular monitoring and management of these women in the peripartum period could have immediate and long-term health benefits requires further evaluation.

**GRAPHIC ABSTRACT:** 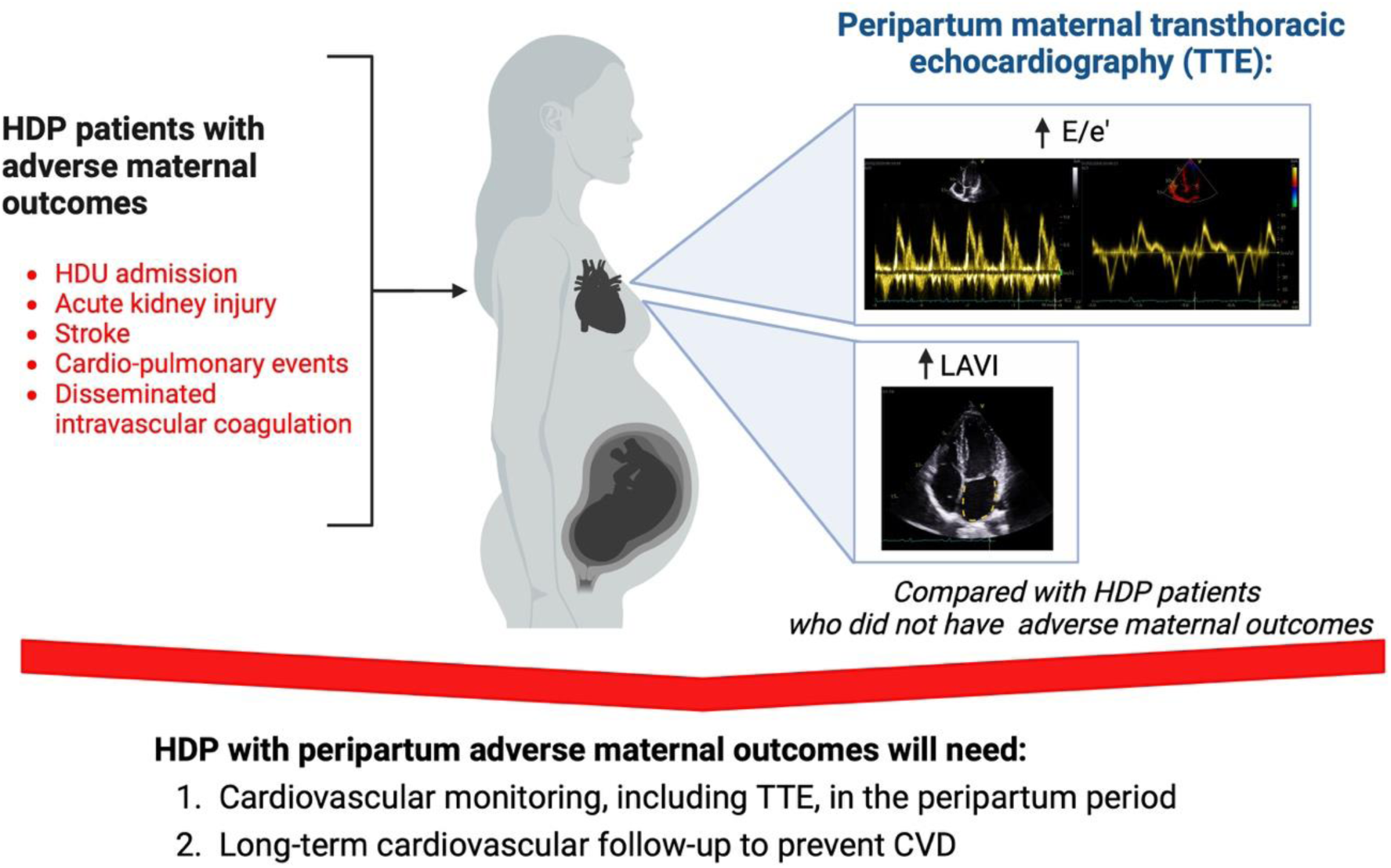

## INTRODUCTION

Hypertensive disorders of pregnancy (HDP), including pre-eclampsia, gestational hypertension and chronic hypertension, are significant contributors to maternal and perinatal morbidity and mortality, affecting approximately 5-10% of pregnancies with an increasing trend globally.^1,2^ In particular, between 5 and 20% of women who develop HDP early in pregnancy can face life-threatening medical complications, such as cerebrovascular hemorrhage, pulmonary edema, acute kidney injury (AKI), liver rupture, and eclampsia.^3^ Worldwide, 10 to 15% of direct maternal deaths are associated with HDP, and mortality is higher in patients with heart disease.^4,5^ Although the involvement of the maternal cardiovascular system in HDP patients has been extensively investigated, to date, no clinical guidelines have formally recommended the assessment of maternal hemodynamic or cardiac function during pregnancy.^6,7^

Transthoracic echocardiography (TTE) can provide valuable insights into left ventricular morphology and function in pregnant women. Several echocardiographic studies have highlighted adverse left ventricular remodeling as well as diastolic and systolic dysfunction in pregnancies complicated by hypertension, with a dose-response relationship in echocardiographic findings across the HDP disease spectrum. ^8–10^ However, few studies have determined whether the development of peri-partum adverse maternal outcomes during HDP are associated with additional significant impacts on the maternal cardiovascular system that may need targeted treatment.^11,12^ In addition, given the potentially preventable long-term impact of HDP on the cardiovascular system,^13^ understanding the peripartum cardiac burden in these high-risk patients could help determine whether enhanced cardiac monitoring and management may also be beneficial post-partum.

## METHODS

This prospective observational study was conducted at St George’s University Hospital NHS Foundation Trust between February 2019 and August 2021. The study enrolled women with HDP undergoing cardiovascular assessment during the peripartum and postpartum periods to investigate the ability to predict persistent postnatal hypertension. The primary results of this study have been previously published.^14–16^ Ethical approval was obtained from the Local Ethics Committee (19/LO/0794), and all participants provided written informed consent. This analysis focused on data from women who underwent TTE either before delivery or within one week postpartum (peripartum period). Median (IQR) days differences between TTE and delivery were 6 (3-14) days before delivery and 2 (1-3) days after delivery. Data from before and shortly after delivery were pooled because our previous findings showed no significant changes in cardiac geometry or function when comparing paired TTE data performed before delivery with data within one week post-delivery.^16^

### Study population

The HDP cohort included women with singleton pregnancies complicated by HDP, such as chronic hypertension, preeclampsia or gestational hypertension, recruited consecutively from the Maternity Department. HDP types were diagnosed according to the criteria established by the International Society for the Study of Hypertension in Pregnancy.^17^ HELLP syndrome was defined when the following criteria were met: liver transaminases >70 IU/L, or twice the upper limit of normal concentration, platelet <100 x 10⁹/L, hemolytic signs like elevated total bilirubin, elevated LDH, and schistocytes on a peripheral blood smear. Hypertension in HDP pregnancies was managed per NICE guidelines using labetalol, nifedipine, or methyldopa based on clinical preferences.^18^ Pregnancies affected by genetic syndromes, major fetal abnormalities, or known cardiac conditions were excluded. Data about pregnancy and delivery outcomes were collected from patients’ medical records. The last laboratory test results were also collected, including platelet count (10^9/L), creatinine (μmol/L), alanine aminotransferase (ALT, U/L), and urinary protein to creatinine ratio (PCR, mg/mmol) performed before delivery.

### Echocardiography measures

Anthropometric measurements, blood pressure (BP), and maternal TTE were performed. Body mass index (BMI, kg/m²) was calculated as weight (kg) divided by height squared (m²), and body surface area (BSA, m²) was calculated using the formula: 0.007184*height(cm)0.725*weight(kg)0.425. Mean arterial pressure (MAP, mmHg) was calculated as (2*diastolic BP+ systolic BP)/3. TTE was performed at rest in the left lateral decubitus position using a GE Vivid E95 with an M5Sc-D probe (GE Healthcare, Horten, Norway), with analysis conducted using EchoPAC version 203 (GE Healthcare). Measurements adhered to international guidelines.^19–22^ From the parasternal long-axis view, the interventricular septum diameter (IVSd), left ventricular end-diastolic diameter (LVEDd) and left ventricular posterior wall thickness (PWT) were measured. Left ventricular mass (LVM) was calculated using the formula: 0.8*(1.04*(LVEDd+PWT+IVS)^3^−LVEDd^3^)+0.6. LVM index (LVMI, g/m^2^) was obtained by indexing for BSA. Relative wall thickness (RWT) was calculated as RWT=2xPWT/LVEDd. Left atrial volume index (LAVI) was measured at end ventricular systole and indexed for BSA. Left ventricular end-diastolic and end-systolic volumes and ejection fraction (EF, %) were measured using the biplane summation-of-disks method.

Pulsed wave Doppler assessed mitral valve flow, including peak early diastolic (E-wave) velocity, atrial contraction (A-wave) velocity, E/A ratio, and deceleration time. Tissue Doppler imaging measured lateral and septal mitral annulus velocities (e′, m/s). The speckle-tracking analysis provided global longitudinal strain (GLS, %) values from the apical 2-, 3-, and 4-chamber views.

### Maternal and perinatal outcomes

The composite adverse maternal outcome was defined as the occurrence of at least one of the following events: admission to a high dependence unit (HDU) for cardiac monitoring or hourly monitoring; AKI defined as creatinine>100 µmol/L antenatally or >130 µmol/L postnatally or need for dialysis; adverse cardiac events, such as acute coronary syndrome, pulmonary oedema, or peripartum cardiomyopathy; hemorrhagic or ischemic stroke; and disseminated intravascular coagulation (DIC).^23^ The composite adverse perinatal outcome was defined as the occurrence of at least one of the following events: perinatal death including stillbirth, neonatal death and termination of pregnancy because of severe fetal growth restriction (FGR) at pre-viable gestational age, preterm delivery before 34 weeks’ gestation, and FGR defined as birthweight below the 3^rd^ centile.^23,24^

### Statistical Analysis

Continuous variables were expressed as medians with interquartile ranges (IQR), and categorical variables were compared using the Chi-square test. Continuous variables were analysed with the Mann-Whitney U-test. Logistic regression models assessed the association between cardiac parameters and composite maternal adverse outcomes. Multivariate analysis was adjusted for maternal factors (maternal age, ethnicity, BMI, MAP, and pre-existing comorbidities) and the timing of TTE to validate these associations. Ordinal logistic regression was used to explore the association of TTE measurements and severity of maternal adverse outcomes that were classified as absent, one and two or more adverse maternal outcomes. Statistical significance was set at p<0.05. Analyses were conducted using SPSS software (version 27.0, SPSS Inc., Chicago, IL, USA).

## RESULTS

The study cohort included 255 pregnancies complicated by HDP (Table 1). The median (IQR) maternal age was 33.8 (29.9-37.3) years, and the first-trimester median (IQR) BMI was 27.0 (23-1-31.2) kg/m². Women identified themself as non-white ethnicity in 39.6% (101/255) of cases. Pre-existing hypertension and diabetes were present in 3.9% (10/255) and 2.4% (6/255) of cases, respectively. The birth weight centile median (IQR) was 16.1 (0.89-53.5), and the gestational age median (IQR) at delivery was 38.0 (35.9-39.4) weeks. Preeclampsia was diagnosed in 153 out of 255 (60%) HDP cases. There were eight cases of HELLP syndrome. Composite adverse maternal outcomes occurred in 26.7% (68/255), while composite adverse perinatal outcomes in 36.1% (92/255). Table 1 shows the frequency of each adverse maternal and perinatal outcome included. 18.1% (46/255) of HDP patients underwent TTE within one week of delivery.

**Table 1.** Description of HDP cohort.

| **Variables** | **Total HDP (N=255)** |
| --- | --- |
| Age (year) | 33.8 (29.9-37.3) |
| 1<sup>st</sup> trimester BMI (kg/m<sup>2</sup>) | 27.0 (23.1-31.2) |
| 1<sup>st</sup> trimester MAP | 94.7 (90.3-99.3) |
| Ethnicity |  |
| Caucasian | 156 (60.4) |
| Afro-Caribbean | 48 (18.8) |
| Asian | 40 (15.7) |
| Mixed/other | 13 (5.1) |
| Smoker | 27 (10.6) |
| IVF | 16 (6.3) |
| Nulliparous | 167 (65.5) |
| Pre-existing CHTN | 10 (3.9) |
| Pre-existing DM | 6 (2.4) |
| High risk of preterm preeclampsia | 70 (34.8) |
| Gestational age at delivery (weeks) | 38.0 (35.9-39.4) |
| Birthweight centile | 16.1 (0.89-53.5) |
| Antihypertensive treatments |  |
| Labetalol | 50 (19.6) |
| Nifedipine | 77 (30.2) |
| Labetalol, nifedipine | 50 (19.6) |
| Methyldopa | 13 (5.1) |
| Labetalol, methyldopa | 1 (0.4) |
| Nifedipine, methyldopa | 11 (4.3) |
| Labetalol, nifedipine, methyldopa | 5 (2.0) |
| **Composite maternal outcome:** | 68 (26.7) |
| • HDU admission | 54 (21.2) |
| • Acute kidney injury | 21 (8.2) |
| • Stroke | 1 (0.4) |
| • Peripartum cardiomyopathy | 1 (0.4) |
| • Pulmonary oedema | 1 (0.4) |
| • Disseminated intravascular coagulation | 1 (0.4) |
| **Composite adverse perinatal outcome:** | 92 (36.1) |
| - Perinatal death | 4 (1.6) |
| - Preterm birth <34 weeks | 37 (14.5) |
| - SGA <3<sup>rd</sup> centile | 86 (33.7) |

Data are expressed as n (%) or median (IQR). HDP hypertensive disorders of pregnancy, BMI body mass index, MAP mean arterial pressure, CHTN chronic
hypertension, DM diabetes mellitus, HDU high dependency unit, SGA small for gestational age.

Women who had composite adverse outcomes delivered significantly earlier (gestational age 35.79 [31.6-38.3] weeks) and had smaller babies (birthweight centile 4.7 [0.1-39.3]) compared to those who did not (gestational age 38.6 [37.1-39.4] weeks, p<0.001; birthweight centile 21.0 [2.3-60.7], p=0.003). They also showed significantly lower platelets and higher creatinine, ALT and PCR compared to women who did not develop adverse maternal outcomes, as expected (Table 2). The univariate analysis, presented in Table 2, highlighted two echocardiographic parameters significantly associated with adverse maternal outcomes in HDP pregnancies. Women with adverse maternal outcomes had higher LAVI (28.8 [23.4-32.3] ml/m^2^vs. 26.6 [22.2-30.9] ml/m^2^, p=0.045) and E/e′ ratio (7.80 [6.59-9.23] vs. 7.00 [5.91-8.08], p=0.002) compared to those without adverse outcomes. These associations remained significant also after adjusting for maternal factors before TTE (maternal age, non-white ethnicity, 1^st^ trimester BMI, 1^st^ trimester MAP and pre-existing co-morbidities) and at the time of TTE (BMI, MAP and timing of TTE including gestational age or days postpartum if TTE was done after delivery), as shown in Table 3. While 44 patients (17.3%) experienced only one adverse outcome, 29 patients (11.4%) had two or more adverse maternal outcomes. In the ordinal regression analysis, E/e’ ratio remained significant (OR 1.28, 95% CI 1.10-1.49, p=0.001), while LAVI was not (OR 1.03, 95% CI 0.98-1.07, p=0.250). As shown in Table 4, none of the echocardiographic parameters was associated with composite adverse fetal or neonatal outcomes.

**Table 2.** Univariate analysis for the association of maternal and cardiac characteristics with adverse maternal outcomes in the HDP cohort.

| Variables | Adverse maternal outcomes (N=68) | No adverse maternal outcomes (N= 187) | P value |
| --- | --- | --- | --- |
| **Maternal characteristics** |  |  |  |
| Age (years) | 33.2 (28.8-36.7) | 34.1 (30.3-37.5) | 0.329 |
| 1<sup>st</sup> trimester BMI (kg/m<sup>2</sup>) | 26.7 (23.0-30.5) | 27.2 (23.4-31.5) | 0.302 |
| 1<sup>st</sup> trimester MAP (mmHg) | 93.0 (89.0-99.3) | 95.3 (90.7-99.3) | 0.517 |
| Non-white Ethnicity | 32 (47.1) | 69 (36.9) | 0.142 |
| 1<sup>st</sup> trimester high risk preterm pre-eclampsia | 17 (37.8) | 53 (34.0) | 0.637 |
| Pre-existing CHTN | 2 (2.9) | 8 (4.3) | 1.000 |
| Pre-existing DM | 2 (2.9) | 4 (2.1) | 0.659 |
| Gestational age at diagnosis | 34.0 (30.0-37.0) | 36.4 (34.0-38.0) | **0.006** |
| **Laboratory parameters** |  |  |  |
| Platelet count (10<sup>9</sup>/L) | 179.5 (130.0-214.0) | 210.0 (172.0-250.0) | **<0.001** |
| Creatinine (μmol/L) | 71.0 (62.0-87.0) | 61.0 (52.0-71.0) | **<0.001** |
| ALT (U/L) | 26.0 (17.0-62.0) | 19.0 (14.0-28.0) | **<0.001** |
| PCR (mg/ mmol) | 133.8 (55.0-459.8) | 37.2 (13.6-94.9) | **<0.001** |
| **TTE parameters** |  |  |  |
| BMI (kg/m<sup>2</sup>) | 30.8 (26.6-33.6) | 31.3 (27.9-35.6) | 0.120 |
| MAP (mmHg) | 104.7 (99.7-108.8) | 103.7 (98.7-108.7) | 0.211 |
| LVMI (g/m<sup>2</sup>) | 78.3 (69.0-92.2) | 77.03 (66.3-86.2) | 0.167 |
| RWT | 0.43 (0.36-0.47) | 0.43 (0.36-0.48) | 0.528 |
| LAVI (ml/m<sup>2</sup>) | 28.8 (23.4-32.3) | 26.6 (22.2-30.9) | **0.045** |
| Lateral e'(m/s) | 0.13 (0.10-0.14) | 0.12 (0.11-0.15) | 0.330 |
| Septal e'(m/s) | 0.09 (0.08-0.12) | 0.10 (0.08-0.12) | 0.364 |
| E/e' | 7.80 (6.59-9.23) | 7.00 (5.91-8.08) | **0.002** |
| EF (%) | 59.0 (57.00-61.0) | 58.0 (55.0-61.0) | 0.152 |
| GLS (%) | -16.3 (-17.8--14.5) | -16.15 (-17.9--14.5) | 0.852 |

Data are expressed as n (%) or median (IQR). HDP hypertensive disorders of pregnancy, BMI body mass index, MAP mean arterial pressure, CHTN chronic hypertension, DM diabetes mellitus, TTE transthoracic echocardiography, LVMI left ventricular mass index, RWT relative wall thickness, LAVI left atrial volume index, EF ejection fraction, GLS global longitudinal strain.

**Table 3.** Multivariable analysis for the association of maternal and cardiac characteristics with adverse maternal outcomes in the HDP cohort.

| Variables | OR (95% CI) | P value | OR<sup>a</sup> (95% CI) | P value | OR<sup>b</sup> (95% CI) | P value |
| --- | --- | --- | --- | --- | --- | --- |
| LAVI (ml/m<sup>2</sup>) | 1.05 (1.01-1.10) | 0.024 | 1.07 (1.02-1.12) | 0.010 | 1.09 (1.02-1.16) | 0.009 |
| E/e' | 1.28 (1.10-1.49) | 0.002 | 1.29 (1.10-1.52) | 0.002 | 1.25 (1.04-1.49) | 0.018 |

<sup>a</sup> adjusted for maternal age, non-white ethnicity, 1<sup>st</sup> trimester BMI, 1<sup>st</sup> trimester MAP and pre-existing co-morbidities
<sup>b</sup> adjusted for BMI, MAP and timing of echocardiography (gestational age, before or after delivery)
OR odd ratio, 95% CI confidence interval, LAVI left atrial volume index.

**Table 4.** Univariate analysis for the association of maternal and cardiac characteristics with composite adverse perinatal outcomes in the HDP cohort.

| Variables | Adverse perinatal outcomes (N=92) | No adverse maternal outcomes (N= 163) | P value |
| --- | --- | --- | --- |
| **Maternal characteristics** |  |  |  |
| Age (years) | 33.8 (29.8-36.5) | 33.99 30.22 39.12 | 0.438 |
| 1<sup>st</sup> trimester BMI (kg/m<sup>2</sup>) | 27.2 (22.9-31.1) | 26.10 23.40 31.65 | 0.881 |
| 1<sup>st</sup> trimester MAP (mmHg) | 95.7 (92.2-99.3) | 92.33 88.33 99.00 | **0.005** |
| Non-white Ethnicity | 47 (51.1) | 54 (33.1) | **0.005** |
| 1<sup>st</sup> trimester high risk preterm pre-eclampsia | 27/59 (45.8) | 43/142 (30.3) | **0.036** |
| Pre-existing CHTN | 4 (4.3) | 6 (3.7) | 0.751 |
| Pre-existing DM | 2 (2.2) | 4 (2.5) | 1.000 |
| Gestational age at diagnosis | 32.2 (27.0-35.2) | 37 (36.0-39.0) | **<0.001** |
| **Laboratory parameters** |  |  |  |
| Platelet count (10<sup>9</sup>/L) | 196.0 (145.5-231.5) | 209.0 (168.5-242.5) | **0.042** |
| Creatinine (μmol/L) | 64.0 (56.0-78.5) | 62.0 (52.0-73.0) | 0.195 |
| ALT (U/L) | 24.5 (18.0-53.0) | 18.5 (14.0-27.0) | **<0.001** |
| PCR (mg/ mmol) | 97.0 (41.0-313.4) | 36.4 (14.5-87.8) | **<0.001** |
| **TTE parameters** |  |  |  |
| BMI (kg/m<sup>2</sup>) | 30.4 (26.3-34.2) | 31.6 (28.2-35.6) | **0.044** |
| MAP (mmHg) | 103.5 (99.7-109.3) | 103.7 (98.7-108.0) | 0.438 |
| LVMI (g/m<sup>2</sup>) | 75.5 (67.4-87.2) | 77.4 (67.8-88.1) | 0.769 |
| RWT | 0.43 (0.36-0.49) | 0.42 (0.35-0.47) | 0.271 |
| LAVI (ml/m<sup>2</sup>) | 27.0 (22.9-31.8) | 27.0 (22.3-31.1) | 0.834 |
| Lateral e'(m/s) | 0.13 (0.11-0.15) | 0.12 (0.11-0.15) | 0.770 |
| Septal e'(m/s) | 0.10 (0.08-0.12) | 0.10 (0.08-0.12) | 0.529 |
| E/e' | 7.3 (6.0- 8.4) | 7.2 (6.2-8.4) | 0.647 |
| EF (%) | 58.0 (56.0-61.0) | 58.0 (56.0-61.0) | 0.925 |
| GLS (%) | -16.2 (-17.9-14.4) | -16.3 (-17.9--14.5) | 0.929 |

Data are expressed as n (%) or median (IQR). HDP hypertensive disorders of pregnancy, BMI body mass index, MAP mean arterial pressure, CHTN chronic hypertension, DM diabetes mellitus, TTE transthoracic echocardiography, LVMI left ventricular mass index, RWT relative wall thickness, LAVI left atrial volume index, EF ejection fraction, GLS global longitudinal strain.

## DISCUSSION

Pregnancies complicated by HDP and adverse maternal outcomes demonstrated significantly higher LAVI and E/e′ values, underscoring the association between diastolic dysfunction and maternal morbidity in this high-risk population. In particular, E/e’ was also associated with a more severe phenotype of adverse maternal outcomes. In contrast, echocardiographic parameters do not seem to be related to adverse perinatal outcomes. These findings indicate that HDP patients who develop adverse maternal outcomes could benefit from closer cardiac monitoring during the peripartum and optimal postnatal management to improve their cardiovascular outcomes.^25^

The findings we observed are physiologically plausible, as left ventricular remodeling of cardiac structure and impaired diastolic function are common features in pregnancies affected by HDP. ^9,10^ There is a clear dose-response relationship between the severity of HDP and echocardiographic abnormalities, reinforcing this study’s results.^11^ Alhuneafat and colleagues demonstrated that women with chronic hypertension and superimposed preeclampsia exhibited the most severe abnormalities, including significantly elevated LVMI and E/e’ ratio; women with preeclampsia showed intermediate changes, and those with gestational hypertension exhibited the mildest changes, often not significantly different from normotensive controls after adjustment for confounders.^11^ However, few prior studies have linked cardiac dysfunction detected during pregnancies complicated by HDP with adverse maternal outcomes. In a prospective observational study, Vaught et al. found that women with preeclampsia with severe features demonstrated higher right ventricular systolic pressures (31.0±7.9 mmHg vs 22.5±6.1 mm Hg, p <0.001), abnormal left ventricular diastolic parameters (septal E/e’ ratio: 10.8 ±2.8 vs 7.4±1.6, left atrial area size: 20.1± 3.8 cm^2^ vs. 17.3±2.9 cm^2^), and increased left-sided cardiac remodeling (PWT: 1.0 cm [0.9 to 1.1 cm] vs 0.8 cm [0.7 to 0.9 cm]) compared to healthy pregnant controls. Notably, within the preeclamptic group, six women (9.5%) developed peripartum pulmonary oedema, further emphasizing the association between cardiac dysfunction and adverse outcomes.^12^

Our study demonstrated a more significant cardiovascular impact, particularly on diastolic function, in HDP pregnancies with maternal adverse outcomes compared to those without such complications. This highlights a potential need for close cardiac monitoring, including TTE, in this subgroup of HDP patients during the peripartum period, as they may be more likely to require cardiac support. Whether these changes are present prior to the onset of adverse maternal outcomes is also of interest as, if so, echocardiographic findings in HDP might assist in the early identification of women at high risk of developing maternal complications ^26–28^ At present, risk in hypertensive pregnancies may be assessed using the full Preeclampsia Integrated Estimate of Risk (PIERS) model. This is a validated predictive tool designed to estimate the risk of adverse maternal outcomes in women with preeclampsia, including eclampsia, stroke, pulmonary oedema, renal failure, liver dysfunction, DIC, placental abruption, and maternal death within 48 hours or 7 days.^29^ It integrates clinical parameters, such as gestational age and symptoms (chest pain and dyspnea), oxygen saturation, and laboratory parameters, including liver function, creatinine and platelet count. After external validation, it demonstrated moderate predictive accuracy, with a pooled AUC of 0.78 (95% CI 0.71-0.86) for outcomes within 48 hours and 0.75 (95% CI 0.69–0.82) within 7 days, with better performance in high-income countries than low- and middle-income countries.^26^ Whether inclusion of echocardiographic parameters could improve interest required further work.

Evidence shows that women with HDP who exhibit worse cardiac remodeling and diastolic dysfunction are at a significantly higher risk of developing chronic hypertension as early as 4–6 months postpartum.^14^ Chronic hypertension, in turn, is the primary risk factor for subsequent cardiovascular complications, such as heart failure and myocardial infarction, in the female population.^13,30^ The Physician Optimized Postpartum Hypertension Treatment (POP-HT) trial has provided compelling evidence of the importance of structured interventions during the postpartum period. In this trial, 220 women with HDP were randomized to either physician-guided self-monitoring of blood pressure with optimized antihypertensive medication titration or standard postpartum care. The intervention group showed significantly better blood pressure control at 6–9 months postpartum.^25^ Moreover, echocardiographic and cardiac magnetic resonance imaging revealed that the intervention group experienced improved cardiac remodeling, including reductions in left ventricular mass, end-diastolic and end-systolic volumes, and relative wall thickness, alongside improvements in left and right ventricular ejection fractions.^31^ These findings underscore the critical role of early and intensive postpartum management in improving outcomes for women with HDP. Whether the more extreme cardiac changes we observed in those who had adverse maternal outcomes in the current study persist post-partum, and can be reversed by improved post-partum care, will be of interest.

The strengths of this study include its prospective design, rigorous echocardiographic assessments, and a well-defined cohort of women with HDP with ethnic diversity. However, there are several limitations. The study was conducted at a single center, potentially limiting generalizability to other populations. Furthermore, although this study is a sub-analysis of a cohort initially designed to assess postpartum outcomes, the study is not adequately powered to evaluate adverse maternal outcomes as a primary predictor of future endpoints and our analysis was limited to a case control analysis.^14–16^ The small sample size, particularly for severe adverse events, also restricted the statistical power to detect additional significant associations or associations with specific individual adverse outcomes. Lastly, the lack of standardized management protocols across participants may have introduced variability in clinical outcomes and intervention bias. Despite these limitations, the findings underscore the potential utility of integrating TTE into clinical practice for managing HDP pregnancies in the peripartum and postpartum.

### Perspectives

This study highlights a significant potential role for echocardiographic parameters, particularly LAVI and E/e’ ratio, to identify women at risk of adverse maternal outcomes in pregnancies complicated by HDP. These findings advocate for further work to understand whether the integration of cardiac assessments into routine peripartum clinical management for HDP with adverse maternal outcomes could allow for targeted monitoring and timely interventions to mitigate peripartum adverse maternal outcomes. Echocardiographic findings obtained during the peripartum could inform targeted postnatal interventions to improve cardiovascular health and reduce long-term risk. Prospective research is essential to validate these findings in larger cohorts designed to evaluate maternal complications as primary endpoints. Moreover, future work should prioritize incorporating echocardiographic markers into predictive tools to determine whether they enhance accuracy and clinical application for the prediction of peripartum maternal complications.

## Data Availability

Data are available on request.

## Acknowledgments

Graphic abstract was created with Biorender.com.

Veronica Giorgione received funding from the European Union’s Horizon 2020 research and innovation programme under the Marie Skłodowska-Curie grant agreement No 765274 (iPLACENTA project).

## Novelty and Relevance

### What Is New?

- This study identifies the independent association of echocardiographic markers, particularly LAVI and E/e′ ratio, with adverse maternal outcomes in pregnancies complicated by hypertensive disorders (HDP).

### What Is Relevant?

- HDP are major contributors to maternal morbidity and mortality worldwide during pregnancy, in addition to the cardiovascular burden after birth.
- HDP pregnancies with maternal complications may require close monitoring because of their impaired cardiovascular function.

### Clinical Implications

- Enhanced peripartum cardiovascular monitoring, including echocardiography, may be required for women with HDP and adverse maternal outcomes, enabling timely interventions to mitigate immediate complications and optimize care during this critical period.
- Adverse maternal outcomes in HDP may signify significant underlying cardiovascular dysfunction, highlighting the need for long-term monitoring and tailored interventions to prevent chronic hypertension and future cardiovascular complications.

